# STUDY OF FACTORS ASSOCIATED WITH PSYCHOSOCIAL WELL-BEING DURING THE COVID-19 PANDEMIC IN SENEGAL

**DOI:** 10.1101/2025.04.15.25325869

**Authors:** Fatou Kasse, Jean Augustin Tine, Adama Faye, Ibrahima Gaye, Amadou Ibra Diallo, Valery Ridde

## Abstract

During the COVID-19 pandemic, the global population was particularly vulnerable to psychological distress. However, the impacts of pandemics on the state of psychosocial well-being in Africa remain insufficiently studied, particularly in Senegal. Our study aimed to fill this gap by exploring these determinants. This was a cross-sectional, descriptive and analytical study representative of the population of Senegalese people aged 18 and over. 813 individuals were collected by telephone call based on the random dialing method after a marginal quota survey stratified by sex, age and region during the period from June 11 to July 10, 2020. The state of well-being was assessed using the WHO well-being index. The determinants were assessed using cumulative ordinal regression well-being modeling with R software version 4.1.0. About half of our population had a moderate level of well-being (48.2%) and about a tenth had a low level (6.6%) including 4 2.6% with a low economic level. The analysis revealed that poor knowledge about the cause of the disease (OR = 1.31; 95% CI [1.09-1.58]), reduced time spent in public places (OR = 1.53; 95% CI [ 1.15-2.04 ]); trust in institutional sources of information (OR = 1.26; 95% CI [ 1.06-1.49 ]) and cancellation or postponement of a social event (OR = 1.34; 95% CI [ 0.96-1.89 ]) are factors related to the decline in psychosocial well-being towards the low level. The study highlights the importance of implementing appropriate communication strategies to strengthen the knowledge of populations at all levels, but also the need to pay particular attention to vulnerable groups and to provide psychosocial support throughout pandemics.

## Introduction

The global social context has been exposed to high stress since the isolation of a new coronavirus in China on December 31, 2019 in the Wuhan province (**1**). The explosion of cases in the Asian continent and then beyond prompted the WHO to declare it a public health emergency of international concern. The disease was then classified as a COVID-19 pandemic on March 11, 2020 by the WHO, which invited all countries to take measures to limit its progression (**2–4**). Following these recommendations, the governments of the affected countries imposed barrier measures by banning public gatherings, physical distancing of individuals, the establishment of curfews and confinement, thus leading to a slowdown in global economic activity and enormous geopolitical and economic consequences (**5**).

All continents were affected and Senegal was the fourth African country to confirm the presence of the virus on its territory in March 2020 (**6**). The increase in cases forced the authorities to declare a state of emergency and to introduce measures to limit its progression. Among other things, they proceeded to close schools and universities, to reorganize working hours in state administrative services, to reduce travel, to prohibit gatherings, to strengthen border controls and to establish a curfew from 8 p.m. to 6 a.m. (**7–10**).

However, studies of previous epidemics such as SARS in 2003 (due to Sars-CoV-1), H1N1 in 2009 (Influenza A virus) and Ebola in 2013 in West Africa had shown that large-scale infectious phenomena could have an impact on mental health (**11,12**). Psychological distress was assessed following the H1N1 influenza epidemic in 2009 and the average rate of psychological distress among health workers was approximately 40% with an average rate of insomnia of 39%. Depression was assessed in seven studies after the SARS epidemic in 2003 with an average rate of approximately 46 % (**13**). However, very few studies have been conducted to assess psychosocial well-being at the population level in a pandemic context.

Psychosocial well-being is a multidimensional concept that encompasses psychological, social and emotional dimensions. Mental health and psychosocial well-being depend not only on a person’s psychological resources but also on the social context in which they find themselves and the environment in which they evolve. These determinants influence each other dynamically and can equally threaten or protect the person’s state of mental health and well-being (**14**). During the pandemic, factors such as financial insecurity, social isolation, and health uncertainty can potentially contribute to a deterioration of this balance. The long-term effects of the pandemic and the measures to manage its spread were therefore a dual global health issue. Psychosocial support for the population should be thought of collectively for the population because mental health and psychosocial well-being must be an integral part of the global health approach and public health strategies.

Despite growing attention to the psychosocial impacts of the pandemic, challenges persist in identifying the specific factors that positively or negatively influence them in diverse socio-cultural and economic contexts.

The state of psychosocial well-being of the Senegalese population during the epidemic has, to our knowledge, never been studied. This research aims to identify the factors positively and negatively associated with psychosocial well-being related to the COVID-19 pandemic. It will thus contribute to the development of intervention strategies and public policies adapted to strengthen the resilience of populations in the face of future crises and to face new challenges of mental and social health. We tried to answer these different questions: What are the main socioeconomic factors influencing psychosocial well-being during the COVID-19 pandemic? How have containment and social distancing policies affected the mental health and psychosocial well-being of individuals?

## MATERIALS AND METHODS

### Study framework

Senegal is a West African country with 14 administrative regions. The population is estimated at 16,209,125 in 2019 compared to 15,726,056 inhabitants in 2018 for an intercensal growth rate of 2.5%. The female population (50.22%) is slightly larger than the male population (49.78%) in 2019 (15).

The number of mobile phone lines was 17,880,594 at the end of 2019, an increase of 7.97% compared to 2018. The mobile phone penetration rate follows the trend of the number of lines, it is 110.31% in 2019, which gives a ratio of telephone numbers per person equal to 1.1 (**16**).

### Type of study

This was a nationwide, cross-sectional, descriptive and analytical study conducted remotely via telephone call from a call center. Data were collected from June to July 2020.

### Study population

The study targeted adult populations in Senegal, aged 18 and over, with a mobile phone capable of answering the questions. A proportional distribution of participants was made according to age, sex and region in order to ensure demographic representativeness.

### Sampling

The sampling process was designed to ensure representativeness of the adult populations of Senegal and Benin who own a mobile phone. The sample size was determined using simulations based on the following formula (**17,18**):

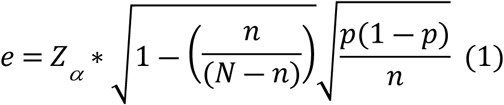

▪ N = Parent population size
▪ n = Sample size
▪ p = Expected proportion in the population
▪ α= Confidence level
▪ *Z_α_*= Value read from the standard normal distribution table

Simulations showed that a sample of 1,000 individuals provides an accuracy of about 3% when the size of the parent population exceeds 100,000. A marginal quota sample was therefore carried out (**19**). The aim is to create a sample that is identical in terms of properties to the parent population. The following variables were used to define the quotas: age, sex and region (**20**) according to the population reference of the last general census in 2013 (**21**). This method is relevant in emergency situations such as the COVID-19 pandemic with sample sizes of less than 3,000 (**22**). A relevant choice of quotas can reduce the variance of the estimate and the amplitude of its confidence interval. When carried out rigorously, the quota sampling method can be as accurate as random sampling or even more so if the sample size is small (**22**). The survey questionnaire was administered to a final sample of 813 individuals. The deviation from the target sample is linked to non-responses from the targeted individuals or to collection constraints (network problems, unavailability, refusals, etc.).

### Operational definition of variables

Well-being comes from the field of research on quality of life. The WHO has constructed a short questionnaire known as WHO-5 which covers 5 topics related to the state of well-being. ***(Table I)***

**Table I:** WHO Well-being Index (1999)

| Over the past two weeks | All the time | Most of the time | More than half the time | Less than half the time | From time to time | Never |
| --- | --- | --- | --- | --- | --- | --- |
| **1. I felt good and in a good mood** | 5 | 4 | 3 | 2 | 1 | 0 |
| **2. I felt calm and peaceful** | 5 | 4 | 3 | 2 | 1 | 0 |
| **3. I felt energetic and vigorous.** | 5 | 4 | 3 | 2 | 1 | 0 |
| **4. I woke up feeling refreshed and alert.** | 5 | 4 | 3 | 2 | 1 | 0 |
| **5. My daily life has been filled with interesting things** | 5 | 4 | 3 | 2 | 1 | 0 |

Composed of 5 items, the questionnaire asks about what people have felt over the past two weeks and understood. There are six response modalities rated from 5 to 0. An overall score is obtained by adding the responses to the 5 items and varies from 0 to 25. Well-being was considered good when the respondent had a score above 20, moderate between 14 and 20 and low when the score was less than or equal to 13. The Cronbach alpha of our well-being scale is greater than 0.7 and shows us a good adequacy of the scale. This short questionnaire was integrated into our questionnaire.

#### Acceptability of the government’s 4 measures

To deal with the pandemic, the Senegalese government had taken measures against the population including a curfew, a ban on travel, and the closure of markets and places of worship. (**7–10**). Each was measured by 7 items that allowed us to have a score ranging from 0 to 7. In our framework, we considered that a measure was respected when the respondent had a score greater than or equal to 6. Compliance with a measure was coded 1 and non-compliance 0. We considered that a respondent accepted the 4 measures when he had a score of 1 in all 4 measures.

#### Acceptability of management of severe cases and contacts at home

It was measured by a 5-point Likert scale. We considered the responses “strongly agree” and “agree” as “yes” and the responses “no” and “don’t know” as “no”.

#### Attitudes and practices related to concern about the epidemic and trust in the government to deal with it

Concern about the COVID-19 pandemic was measured by a scale from 0 to 10 ranging from not at all worried to completely allowing for a broadening in the expression of responses. In our framework, we consider that the individual was worried when he had the maximum score. Like concern, the same scale from 0 to 10 made it possible to assess confidence in the government on the efficient management of the pandemic (0 not at all confident and 10 completely agree and confident with the government management). As with concern, the individual was considered to have confidence in the government when he had the maximum score.

### Data collection and management

The questionnaire was constructed from the WHO guidelines on COVID-19 (**19**), based mainly on two theoretical models of intention regarding health behaviors. On the one hand, the theory of planned behavior (TPB) which postulates that attitudes, subjective norms and perceived behavioral control are the determining factors of an individual’s actual health behavior. On the other hand, the Health Belief Model (HBM) which maintains that the perception of susceptibility to a particular health problem, the perception of the severity of the disease; the belief in the effectiveness of the new behavior; the incentive to action, the perceived benefits of preventive action, the perceived barriers to action (perceived risks; perceived difficulties etc.), represent key predictors of health behavior.

The constructed questionnaire, in electronic Open Data Collet (ODK) format, was integrated into a networked computer program with an internet and telephone component. The program selects unique telephone numbers (n= 30,603) randomly according to the country-wide numbering plan using the *Random Digit Dialing* (RDD) method. (**23**). A second computer program sent an SMS to the previous list to provide information about the project (including ethical issues) and to warn subscribers that they were likely to be called. This program identified valid numbers by the delivery status of the SMS. This made it possible to deduce a new list of numbers supposedly valid (n = 10,931). Then, this list is injected into a Reactive Auto Dialer (RAD) call automation to trigger calls automatically and in an optimized way (n = 6,576) (**24**). At this point, the person was connected to a female interviewer (n= 1441) based in Dakar who explained the research and asked for consent to participate. Before data sharing, all identifying information such as geographic location, names and telephone numbers were removed to maximize respondent confidentiality. Only domain identification codes were retained in the electronic data files.

The collection was carried out by 5 female investigators speaking the main national languages and French after three days of training on the process and content of the survey.

### Data analysis

Data were collected and analyzed using R software version 4.1.0.

The descriptive analysis was done by calculating or determining the position parameters (frequencies for categorical variables and average for quantitative variables) and the dispersion parameters (standard deviation). The bivariate analysis was done by crossing well-being and the other variables to translate the concerns formulated in the research objectives of the associated factors. The Chi 2 test was used considering a difference as significant when the p is less than 0.05. The Student test was also performed considering the same alpha risk to compare the means. A modeling of the state of well-being was carried out in order to search for and determine the factors associated with the low level of well-being. A cumulative ordinal logistic regression model with three levels of psychosocial well-being (good, moderate and low) was obtained using the "step-by-step descending" method until the most parsimonious model was obtained. The reliability of the Likert scale of well-being was tested and the Cronbach alpha was equal to 0.85 and showed us a good reliability.

### Ethical considerations

The research received approval from the National Committee on Health Research Ethics of Senegal (SEN/20/23). To this end, the participant was informed of the objective, methodology and expected results of the study. The respondent was informed that participation is voluntary, anonymous and that he or she can stop the interview at any time.

## RESULTS

### Sample characteristics

Of the 813 individuals surveyed, the average age of the respondents was 34.70 years with a standard deviation of 14.19 years. The median age was 31.00 years. The proportion of individuals over 30 years old was 50.8%. Male individuals represented 54.6% of our study and 61.4% were married (499). The level of economic well-being was low in 20.3% of the respondents. The majority of individuals had poor knowledge of the cause of the disease with a proportion of 72.9% and of the signs or 73.2% while 72.6% had a good level of knowledge of the modes of transmission. A proportion of 79.5% of the respondents did not adhere to the 4 government measures (curfew, ban on travel between regions, closure of markets and closure of places of worship). Furthermore, the study showed that the majority of respondents respected the barrier measures to combat contamination; 90.2%, 87.9% and 96.2% of respondents changed their attitudes on respectively the time spent in public places, the modification or reduction of the use of public transport and the wearing of masks. ***(Table II)***

**Table II:**
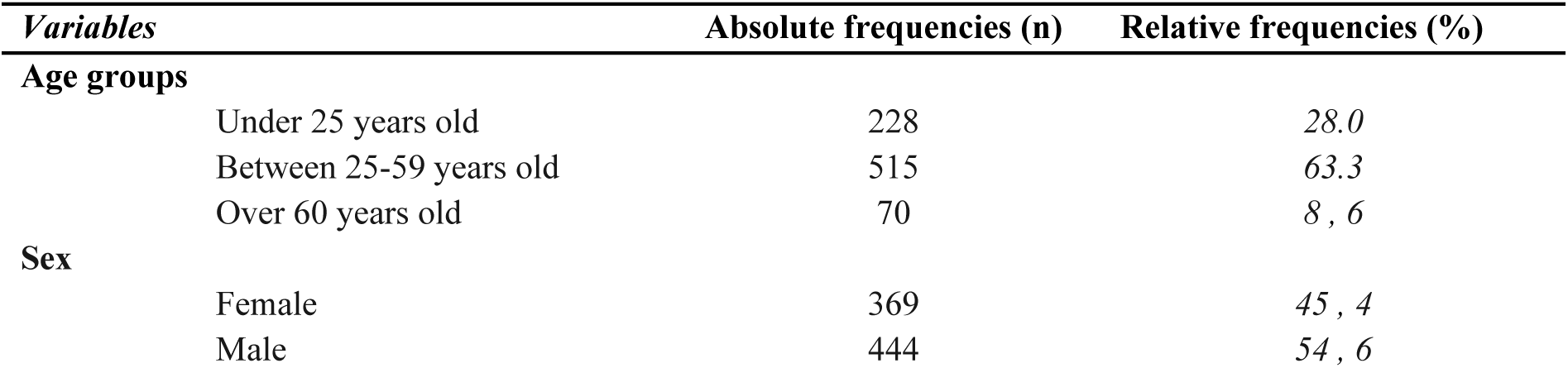
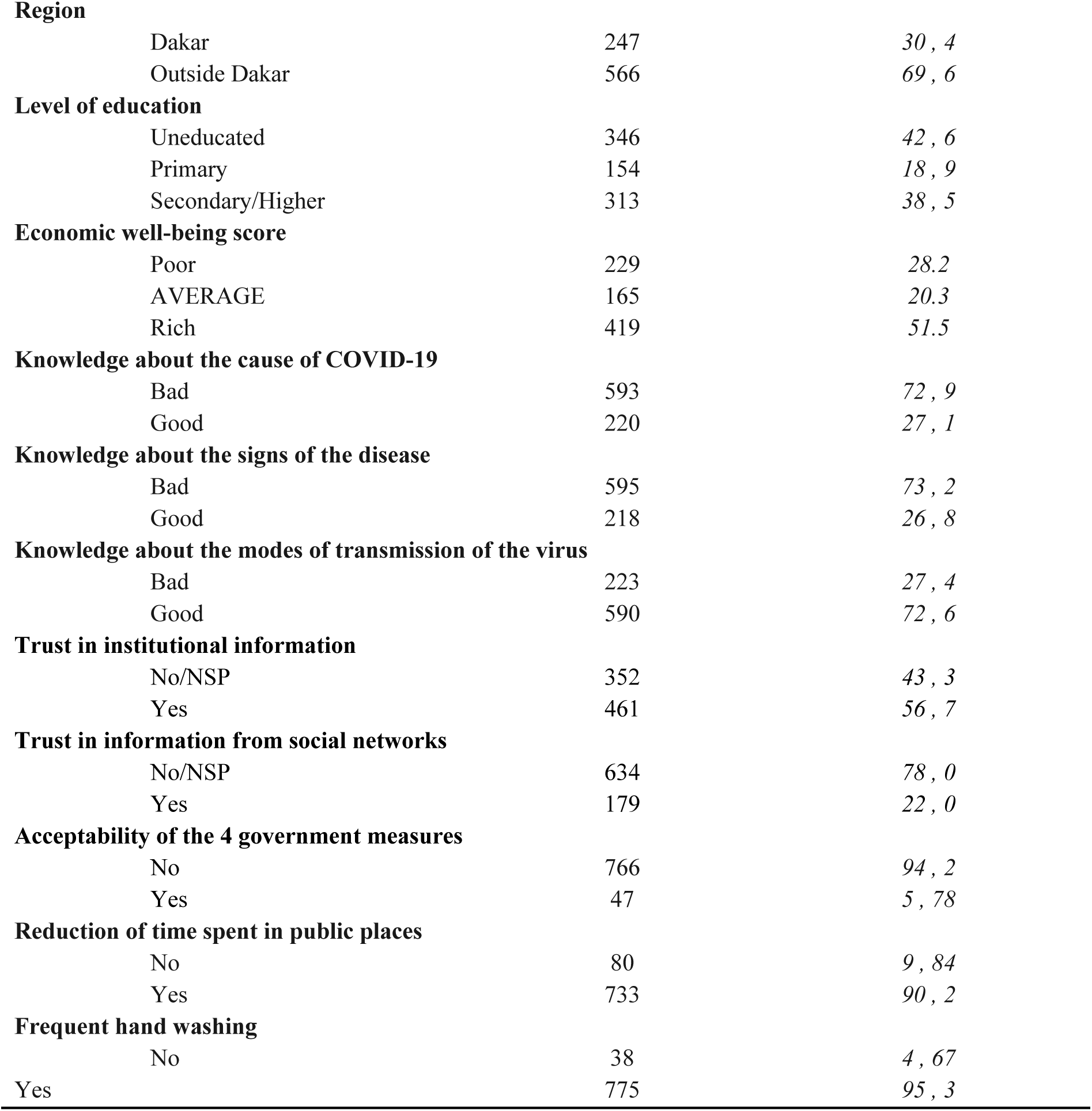
Results of the descriptive analysis.

| *Variables* | *Absolute frequencies (n)* | *Relative frequencies (%)* |
| --- | --- | --- |
| **Age groups** |  |  |
| Under 25 years old | 228 | 28.0 |
| Between 25-59 years old | 515 | 63.3 |
| Over 60 years old | 70 | 8, 6 |
| **Sex** |  |  |
| Female | 369 | 45, 4 |
| Male | 444 | 54, 6 |
| **Region** |  |  |
| Dakar | 247 | 30 , 4 |
| Outside Dakar | 566 | 69 , 6 |
| **Level of education** |  |  |
| Uneducated | 346 | 42 , 6 |
| Primary | 154 | 18 , 9 |
| Secondary/Higher | 313 | 38 , 5 |
| **Economic well-being score** |  |  |
| Poor | 229 | 28.2 |
| AVERAGE | 165 | 20.3 |
| Rich | 419 | 51.5 |
| **Knowledge about the cause of COVID-19** |  |  |
| Bad | 593 | 72 , 9 |
| Good | 220 | 27 , 1 |
| **Knowledge about the signs of the disease** |  |  |
| Bad | 595 | 73 , 2 |
| Good | 218 | 26 , 8 |
| **Knowledge about the modes of transmission of the virus** |  |  |
| Bad | 223 | 27 , 4 |
| Good | 590 | 72 , 6 |
| **Trust in institutional information** |  |  |
| No/NSP | 352 | 43 , 3 |
| Yes | 461 | 56 , 7 |
| **Trust in information from social networks** |  |  |
| No/NSP | 634 | 78 , 0 |
| Yes | 179 | 22 , 0 |
| **Acceptability of the 4 government measures** |  |  |
| No | 766 | 94 , 2 |
| Yes | 47 | 5 , 78 |
| **Reduction of time spent in public places** |  |  |
| No | 80 | 9 , 84 |
| Yes | 733 | 90 , 2 |
| **Frequent hand washing** |  |  |
| No | 38 | 4 , 67 |
| Yes | 775 | 95 , 3 |

### The state of psychosocial well-being of the study population

The proportion of individuals who had a low state of well-being was 6.6% and the majority were in the moderate range with a proportion of 48.2%. ***(Table III)***

**Table III:** Descriptive results of psychosocial well-being.

| *Variables* | *Absolute frequencies (n)* | *Relative frequencies (%)* |
| --- | --- | --- |
| **Psychosocial well-being** |  |  |
| Good | 367 | 45.1 |
| Moderate | 392 | 48.2 |
| Down | 54 | 6.6 |

### Factors related to psychosocial well-being

The study shows that respondents who did not live on the Dakar-Diourbel-Thiès axis had a low state of well-being at 9.4% and those living in this axis at 4.79%. The Dakar-Diourbel-Thiès residential axis is statistically linked to the state of well-being, p-value = 0.043. Individuals who had a low level of well-being did not know the cause of the disease (7.76%) and had a low level of knowledge on the modes of transmission of the virus (5.98%). The state of well-being is statistically linked to knowledge about the cause of the disease and the modes of transmission with a p-value respectively equal to 0.026 and 0.021. The study showed that there is a link between psychosocial well-being and the level of economic well-being (p-value=0.034). It was also found a link with knowledge about the cause of the disease (p-value=0.026) and with the acceptability of the management of simple cases at home (p-value=0.033). A statistical link was also established between psychosocial well-being and the reduction of time spent at work and in public places (p-value=0.009 and 0.036) and the use of hydroalcoholic solutions (p-value=0.031). ***(Table IV)***

**Table IV:** Results of bivariate analysis.

| Variables | STATE OF WELL-BEING |  |  | p-value |
| --- | --- | --- | --- | --- |
|  | Down | Moderate | Good |  |
|  | n(%) |  |  |  |
| Age groups |  |  |  |  |
| 25 - 59 years old | 31 (6 , 02) | 248 (48 , 2) | 236 (45 , 8) | 0.745 |
| < 25 years old | 17 (7 , 46) | 109 (47 , 8) | 102 (44 , 7) |  |
| > 60 years old | 6 (8 , 57) | 35 (50 , 0) | 29 (41 , 4) |  |
| Sex |  |  |  |  |
| Female | 29 (7 , 86) | 180 (48 , 8) | 160 (43 , 4) | 0.363 |
| Male | 25 (5 , 63) | 212 (47 , 7) | 207 (46 , 6) |  |
| Marital status, married |  |  |  |  |
| No | 22 (7 , 01) | 143 (45 , 5) | 149 (47 , 5) | 0.480 |
| Yes | 32 (6 , 41) | 249 (49 , 9) | 218 (43 , 7) |  |
| Residential axis |  |  |  |  |
| Others | 32 (9 , 04) | 171 (48 , 3) | 151 (42 , 7) | 0.043 |
|  | Down | Moderate | Good |  |
|  | n(%) |  |  |  |
| Dakar-Diourbel-Thies | 22 (4 , 79) | 221 (48 , 1) | 216 (47 , 1) |  |
| Quintile |  |  |  |  |
| Poor | 23 (10 , 0) | 117 (51 , 1) | 89 (38 , 9) |  |
| AVERAGE | 9 (5 , 45) | 84 (50 , 9) | 72 (43 , 6) | 0.034 |
| Rich | 22 (5 , 25) | 191 (45 , 6) | 206 (49 , 2) |  |
| Knowledge about the cause of COVID-19 |  |  |  |  |
| Virus | 8 (3 , 64) | 99 (45 , 0) | 113 (51 , 4) |  |
| Other | 46 (7 , 76) | 293 (49 , 4) | 254 (42 , 8) | 0.026 |
| Knowledge about the modes of transmission of the virus |  |  |  |  |
| Down | 24 (10 , 8) | 112 (50 , 2) | 87 (39.0) |  |
| Good | 12 (4 , 15) | 137 (47 , 4) | 140 (48 , 4) | 0.021 |
| AVERAGE | 18 (5 , 98) | 143 (47 , 5) | 140 (46 , 5) |  |
| Acceptability of the 4 government measures |  |  |  |  |
| No | 49 (6 , 40) | 370 (48 , 3) | 347 (45 , 3) |  |
| Yes | 5 (10 , 6) | 22 (46 , 8) | 20 (42 , 6) | 0.477 |
| Support for simple cases at home |  |  |  |  |
| No | 21 (5 , 32) | 208 (52 , 7) | 166 (42 , 0) |  |
| Yes | 33 (7 , 89) | 184 (44 , 0) | 201 (48 , 1) | 0.033 |
| Reduction of working time |  |  |  |  |
| No | 8 (2 , 90) | 138 (50 , 0) | 130 (47 , 1) |  |
| Yes | 46 (8 , 57) | 254 (47 , 3) | 237 (44 , 1) | 0.009 |
| Reduction of time spent in public places |  |  |  |  |
| No | 4 ( 5,00 ) | 29 (36 , 2) | 47 (58 , 8) |  |
| Yes | 50 (6 , 82) | 363 (49 , 5) | 320 (43.7) | 0.036 |
| Frequent hand washing |  |  |  |  |
| No | 3 (7 , 89) | 17 (44 , 7) | 18 (47 , 4) | 0.805 |
| Yes | 51 (6 , 58) | 375 (48 , 4) | 349 ( 45,0 ) |  |

### Modeling psychosocial well-being

The analysis reveals a tendency to go from the good level of well-being to the poor level in individuals who had poor knowledge about the cause of the disease (OR = 1.31; 95% CI [1.09-1.58]), in those who had reduced the time spent in public places (OR = 1.53; 95% CI [ 1.15-2.04 ]); in individuals who had confidence in institutional sources of information (OR = 1.26; 95% CI [ 1.06-1.49 ]) and in those who had cancelled or postponed a social event (OR = 1.34; 95% CI [ 0.96-1.89 ]). Furthermore, a tendency to have a good level of well-being was observed among those who used hydroalcoholic solutions to disinfect their hands (OR = 0.76; 95% CI [ 0.63-0.92 ]) and among those who trusted the information circulating on social networks (OR = 0.79; 95% CI [ 0.64-0.96 ]). ***(Table V)***

**Table V:** Results of the modeling of psychosocial well-being.

|  |  | ORa | 95% CI | p-value |
| --- | --- | --- | --- | --- |
| Well-being level | Good state of well-being/Moderate state of well-being | 1 , 39 | 1 , 01-1 , 64 | 0.065 |
|  | Moderate state of well-being/Low state of well-being | 7 , 36 | 7 , 12-7 , 79 | <0.001 |
| *Independent variables* |  |  |  |  |
| Knowledge about the cause of the disease |  |  |  |  |
|  | Bad | 1 , 31 | 1 , 09-1 , 58 | 0.004 |
|  | Good | Ref | Ref |  |
| Reduction of time spent in public places |  |  |  |  |
|  | Yes | 1.53 | 1 , 15-2 , 04 | 0.003 |
|  | No | Ref | Ref |  |
| Use of hydroalcoholic solution |  |  |  |  |
|  | Yes | 0 , 76 | 0 , 63-0 , 92 | 0.005 |
|  | No | Ref | Ref |  |
| Postponement or cancellation of social events |  |  |  |  |
|  | Yes | 1 , 34 | 0.96-1.89 | 0.087 |
|  | No | Ref | Ref |  |
| Trust in institutional sources |  |  |  |  |
|  | Yes | 1 , 26 | 1 , 06-1 , 49 | 0.008 |
|  | No | Ref | Ref |  |
| Trust in social networks |  |  |  |  |
|  | Yes | 0 , 79 | 0 , 64-0 , 96 | 0.021 |
|  | No | Ref | Ref |  |

## DISCUSSION

Our study population had a majority of moderate psychosocial well-being with a prevalence of 48.2% and 6.6% had a low well-being ***(Table III)***. A study carried out in France in January 2020 showed a state of distress in 5% of the population (**25**). Other studies have also revealed a prevalence of between 29.6% and 31.9% of stress state linked to the pandemic (**26**).

The study showed that individuals under 30 were more likely to have a problematic psychosocial well-being score (51.9%) with a predominance among women (53.7%) ***(Table IV)***. This can be explained by the level of unemployment among people aged 15 or over, which was estimated at 16.9% in the fourth quarter of 2019 in Senegal with a predominance among women (27.6%) (**27**). Furthermore, the study revealed that the majority of low well-being was linked to poverty. Another study carried out in Senegal in July 2020 revealed that 85% of households reported experiencing a drop in their income and 36% had stopped working, 30% of which were for reasons related to covid-19. Nearly eight out of ten households perceived a negative change in their well-being since covid-19 (**28**). The closure of regions, markets, restaurants and non-essential production activities posed a danger to certain segments of the population, especially to families already in vulnerable situations. The pandemic has significantly changed the country’s economic outlook. Indeed, growth in 2020 slowed sharply to an estimated 1.3%. (**29**). The economic difficulties linked to the pandemic mean that despite high concern (54.4%), the majority did not adhere to the 4 government measures (79.5%). At the start of the pandemic, the regions of Dakar, Diourbel and Thiès had recorded in record time the majority of cases across the entire national territory (**30**). The study showed a link between the state of well-being and this axis of residence. A low state of well-being was more observed among those who did not live in this axis. This could be explained by the feeling of upheaval and despair in the face of the increase in cases in other regions which can be subject to anxiety, probably due to a high awareness of the presence of the disease in the territory. According to Fishbein and Ajzen (1975), individuals take into account information from the environment and they always have a minimum of information allowing them to have a precise idea of the situation they are facing. In addition, the rapid spread of the virus has prompted the president to implement a number of measures aimed at reducing gatherings and unnecessary contacts (**8**). These imposed health measures have exposed the population to an unforeseen situation. The study showed that individuals who reduced the time spent at work and in public places (OR = 1.53; 95% CI [ 1.15-2.04 ]) were more likely to have a low state of well-being ***(Table V)***. Indeed, the measures taken by the government as well as by health personnel to reduce the circulation of the virus have required adjustments to the population’s lifestyles. In addition, the limitation of access to normal daily activities and social isolation by banning gatherings can lead to an increase in the level of stress and anxiety. These results are similar to studies carried out in China in 2020 where a fairly high rate of psychological distress and anxiety at 38% and 50.9% respectively had been observed. (**2,3,31–33**). A link has been established between the state of well-being and the trust placed in information emitted by institutional sources and information circulating on social networks. Trust is the basis of a good exchange relationship between two parties, in this case, in the relationship between institutions and their administrators (**32**). As much as networks, individuals’ trust in their institutions facilitates cooperation and coordination for mutual benefit. The data showed that 93.1% trusted institutions, similar results to a study conducted in Cameroon (**33**). In our study, trust in institutional sources increased the risk of having a low level of well-being (OR = 1.26; 95% CI [ 1.06-1.49 ]) Unlike social networks where individuals were less likely to have a low level ***(Table V)***. Regarding information on coronavirus disease, preventive measures, barrier measures issued by the institution are credible information of course (given that they are international in nature) and allow users to be informed about the situation of the pandemic but can be a source of stress and anxiety. Unlike institutional sites, social networks are exchange platforms that allow Internet users and professionals to share information, interact and have conversations about their lives and real facts related to Covid. Sharing spaces that are distinguished by their usefulness (personal, professional, meetings, etc.) that make information accessible and can explain the positive link between well-being and the use of social networks.

## BOUNDARIES

The samples were composed only of national representatives and did not allow for disaggregation by residuals. Only people with a mobile phone were interviewed, thus excluding marginalized populations.

## CONCLUSION

The covid-19 pandemic is much more than a health crisis, it is also an unprecedented socio-economic crisis putting pressure on each of the countries it affects; it has devastating socio-economic, political and psychosocial impacts that will leave deep scars. Studies carried out around the world have indicated a negative impact of the pandemic on the state of psychosocial well-being of populations but few have been done in Africa.

In our study, these various factors were identified as being associated with well-being through the analysis of the results. Cancellation or postponement of social events, reduction of time spent at work, use of hydroalcoholic gel, trust in institutional sources, trust in social networks and knowledge about the causes and modes of transmission of the disease were identified as factors associated with psychosocial well-being.

This study revealed empirical aspects on the impact of the pandemic on the state of well-being of the Senegalese population by drawing inspiration from an integrative WHO model to extract the determinants. Additional studies are needed, especially qualitative, in order to determine the more precise aspects of the determinants of the impact of the pandemic on well-being.

## Data Availability

Les données ont été collectées via ODK collecte puis généré sous format de tableur (Microsoft Excel) Les données démographiques enquêtes, leurs habitude de vie, leurs perceptions, leur acceptabilité par rapport aux mesures restrictives de lutte la pandémie et leurs connaissances sur la maladie sont fournis dans le fichier Excel. L’analyse des données a été faite avec le logiciel R version 4.1.0 disponible uniquement en ligne sur ce lien https://figshare.com/s/75a21943c9a06e9a28eb

## Authors’ contributions

Conceptualization: Fatou Kasse, Jean Augustin Tine, Valery Ridde, Adama Faye.

Data retention: Ibrahima Gaye, Fatou Kasse.

Funding search: Valery Ridde, Adama Faye.

Investigation: Fatou Kasse, Valery Ridde, Adama Faye.

Methodology: Fatou Kasse, Jean Augustin Tine, Adama Faye.

Project administration: Jean Augustin Tine, Valery Ridde, Adama Faye.

Software: Fatou Kasse.

Validation: Fatou Kasse.

Visualization: Fatou Kasse.

Writing – original draft: Fatou Kasse.

Writing – revision and editing: Ibrahima Gaye, Jean Augustin Tine, Adama Faye.

## FINANCING

This research is part of the support program for the African Response to the Epidemic of COVID-19 (ARIACOV) funded by the French Development Agency (AFD). The did not play any role in the design of the study, the collection and analysis of the decision to publish or the preparation of the manuscript.

## REFERENCES

1. World Health Organization. Novel Coronavirus (2019-nCoV) [Internet]. World Health Organization. Geneva, Switzerland; 2020. 2020 [cited 24 Dec 2020]. Situation report - 1 Available at: https://www.who.int/fr/emergencies/diseases/novel-coronavirus-2019

2. Bernard Stoecklin S, Rolland P, Silue Y, Mailles A, Campese C, Simondon A, et al. First cases of coronavirus disease 2019 (COVID-19) in France: surveillance, investigations and control measures. January 2020. Eurosurveillance [Internet]. 13 Feb 2020 [cited 24 Dec 2020];25(6). Available at: https://www.ncbi.nlm.nih.gov/pmc/articles/PMC7029452/

3. Zhong BL, Luo W, Li HM, Zhang QQ, Liu XG, Li WT, et al. Knowledge, attitudes, and practices toward COVID-19 among Chinese residents during the rapid rise of the COVID-19 epidemic: a rapid online cross-sectional survey. Int J Biol Sci. 2020 Mar 15;16(10):1745-52.

4. World Health Organization WHO Director-General’s opening remarks at the media briefing on COVID-19 Geneva, Switzerland; March 2020 [Internet]. [Accessed 9 May 2021]. Available at: https://www.who.int/en/director-general/speeches/detail/who-director-general-s-opening-remarks-at-the-media-briefing-on-covid-1911-march-2020

5. Elie Gerschel, Alejandra Martinez, Isabelle Mejean et al. Propagation of shocks in international value chains: the case of the coronavirus. March 2020. HAL-SHS - Human and Social Sciences [Internet]. [accessed 24 Dec 2020]. Available on: https://halshs.archives-ouvertes.fr/halshs-02515354

6. Gilbert M, Pullano G, Pinotti F, Valdano E, Poletto C, Boëlle PY, et al. Preparedness and vulnerability of African countries to COVID-19 importations: a modelling study. Lancet London Engl. 2020 Mar 14;395(10227):871-7.

7. Government of Senegal. Order No. 007782 of March 13, 2020, temporarily banning demonstrations or gatherings [Internet]. Dakar, Senegal; 2020 [cited May 11, 2021]. Available at: https://www.sec.gouv.sn/arr%C3%AAt%C3%A9-n%C2%B0-007782-du-13-mars-2020-portant-interdiction-provisoire-de-manifestations-ou-rassemblements

8. Government of Senegal. Declaration of a state of emergency in the fight against the coronavirus disease COVID-19 [Internet]. Dakar, Senegal; 2020 [cited 31 Jan 2021]. Message from HE the President of the Republic Macky SALL. Available at: https://www.sec.gouv.sn/actualit%C3%A9/message-de-sem-le-pr%C3%A9sident-de-la-r%C3%A9publique-macky-sall-d%C3%A9claration-d%E2%80%99%C3%A9tat-d%E2%80%99urgence-dans

9. Government of Senegal. Decree No. 2020-875 of March 25, 2020, temporarily reorganizing working hours in state administrative services [Internet]. Dakar, Senegal; 2020 [cited June 4, 2021]. Available at: https://www.sec.gouv.sn/actualit%C3%A9/d%C3%A9cret-n%C2%B0-2020-875-du-25-mars-2020-portant-r%C3%A9am%C3%A9nagement-%C3%A0-titre-provisoire-des-horaires

10. Government of Senegal. Ministerial Order No. 008231 of March 25, 2020 relating to restrictive measures in the land transport sector to combat Covid-19 [Internet]. Dakar, Senegal; 2020 [cited June 4, 2021]. Available at: https://www.sec.gouv.sn/actualit%C3%A9/arr%C3%AAt%C3%A9-minist%C3%A9riel-n%C2%B0-008231-du-25-mars-2020-relatif-aux-mesures-de-restriction-dans-le

11. Angelina OM Chan1 and Chan Yiong Huak. Psychological impact of the 2003 severe acute respiratory syndrome outbreak on health care workers in a medium-sized regional general hospital in Singapore. 2004 May 1;54(3):190-6.

12. Kunitaka M, Ayako KM, Hissei I, Atsushi I, Kentaro M, Noboru K, et al. Psychological impact of the 2009 H1N1 pandemic on general hospital staff in Kobe. January 2010 [Internet]. [cited 22 Jul 2024] Available at: https://onlinelibrary.wiley.com/doi/epdf/10.1111/j.1440-1819.2012.02336.x?src=getftr

13. Vyas KJ, Delaney EM, Webb-Murphy JA, Johnston SL, et al. Psychological impact of deployment in support of the US response to Ebola: a systematic review and meta-analysis of past outbreaks. Military Medicine. 2016 Nov 01;181(11-12):e1515-31.

14. World Health Organization. Mental health risks World Health Organization. Geneva, Switzerland; 2012 [Internet]. [Accessed 9 May 2021]. Available on: https://www.who.int/mental_health/mhgap/risks_to_mental_health_FR_27_08_12.pdf

15. Ministry of Economy, Planning and Cooperation. Senegal: Population of Senegal - Statistics by region in 2019. Dakar, Senegal. [cited June 22, 2021]. National Agency of Statistics and Demography (ANSD) <u>Available at:</u> https://samabac.com/population-du-senegal-les-statistiques-par-region-en-2019-ansd

16. Telecommunications and Postal Regulatory Authority (ARTP). Senegal: Status Report 2019. Dakar, Senegal; June. 2020[Internet]. [Cited 26 Sep 2024] <u>Available at:</u> https://artp.sn/rapport-etat-des-lieux-2019-edition-juin-2020

17. Hogan T, Parent N, Stephensen R. Introduction to psychometrics, 2nd edition. Canada: Chenelière-Education; 2017.

18. Laveault D, Gregoire J. Introduction to test theories in human sciences, 3rd edition. Brussels: De Boeck; 2014.

19. Deville JC. A theory of quota surveys. Survey Methodology, Statistics Canada. 1991;17(2):177–95.

20. Ardilly P. Survey techniques. Paris: Editions Technip; 2006. 704 p.

21. Ministry of Economy, Finance and Planning. Senegal: General Census of Population and Housing, Agriculture and Livestock 2013. Dakar, Senegal; 2013[Internet]. National Agency for Statistics and Demography (ANSD) [cited 30 Jan 2021]. Available at: https://anads.ansd.sn/index.php/catalog/51

22. Riandey B, Blöss-Widmer I. Introduction to surveys for use by the greatest number. https://hal.archives-ouvertes.fr/hal-01272371. 2009;

23. Wolter K, Chowdhury S, Kelly J. Chapter 7. Design, conduct, and analysis of random digit surveys. In: Rao CR, ed. Handbook of Statistics [Internet]. Elsevier; 2009 [cited 21 Jan 2021]. pp. 125-54. (Handbook of Statistics; vol. 29). Available from: http://www.sciencedirect.com/science/article/pii/S0169716108000072

24. What is Auto Dialer. When should you invest in it [Internet]. Imaginesales. 2020 [cited 30 Jan 2021]. Available at: https://www.imaginesales.co/auto-dialer/

25. Tourette-Turgis C, Chollier M. Lifestyle changes and psychosocial impact of COVID-19 confinement. Médecine Mal Métabolics. Dec 2020;S1957255720301164.

26. Salari N, Hosseinian-Far A, Jalali R, Vaisi-Raygani A, Rasoulpoor S, Mohammadi M, et al. Prevalence of stress, anxiety, and depression in the general population during the COVID-19 pandemic: a systematic review and meta-analysis. Glob Health [Internet]. 6 Jul 2020 [Accessed 9 May 2021];16. Available at: https://www.ncbi.nlm.nih.gov/pmc/articles/PMC7338126/

27. National Agency of Statistics and Demography. Senegal: Statistical information on the economic and social situation in the context of the pandemic [Internet]. Dakar; Senegal; 2021. [Accessed July 7, 2021]. Available at: https://satisfaction.ansd.sn/

28. Ministry of Economy and Finance. Senegal: HFMSWE Monthly Report, Monitoring the Impact of Covid-19 on Household Well-being - September 2020. Dakar; Senegal; 2020; Bulletin Number 1 [Internet]. [Accessed July 7, 2021]. Available at: http://www.finances.gouv.sn/communique-de-lansd-suivi-de-limpact-de-la-covid-19-sur-le-bien-etre-des-menages-septembre-2020/

29. World Bank in Senegal. Senegal-Overview[Internet]. Dakar, Senegal; 2020. [Accessed January 30, 2021]. Available at: https://www.banquemondiale.org/fr/country/senegal/overview

30. M. Zeynil-El-Abdine - Ndongo. Analysis of the evolution of the covid 19 pandemic in Senegal-CRES | Consortium for Economic and Social Research [Internet]. March 31, 2020. [Accessed January 30, 2021]. Available at: https://www.cres-sn.org/?s=Analyse+de+l%E2%80%99%C3%A9volution+de+la+pand%C3%A9mie+du+covid+19+au+S%C3%A9n%C3%A9gal&searchsubmit=Search

31. Xiong J, Lipsitz O, Nasri F, Lui LMW, Gill H, Phan L, **et al.** Impact of the COVID-19 pandemic on mental health in the general population: a systematic review. J Affect Disord. 1 Dec 2020;277:55-64.

32. Ayaz CM, Dizman GT, Metan G, Alp A, Unal S. Outpatient management of patients with COVID-19 on home isolation. Infez Med. 2020 Sep 01;28(3): 351–356. [Accessed 17 Jul 2021]. Available at: https://medworm.com/819995194/out-patient-management-of-patients-with-covid-19-on-home-isolation/

33. Kouabénan DR, Cadet B, Hermand D, Sastre MTM. Chapter 12. From beliefs to protective behaviors. De Boeck Supérieur;2007. 143–154. [Accessed 17 Jul 2021]. Available on: https://www.cairn.info/article.php?ID_ARTICLE=DBU_KOUAB_2007_01_0143

